# Understanding the spreading patterns of COVID-19 in UK and its impact on exit strategies

**DOI:** 10.1101/2020.05.18.20105445

**Authors:** Maziar Nekovee

**Affiliations:** School of Engineering and Informatics, University of Sussex, UK

## Abstract

Prior to lockdown the spread of COVID-19 in UK is found to be exponential, with an exponent α=0.207. In case of COVID-19 this spreading behaviour is quantitatively better described with a mobility-driven SIR-SEIR model [2] rather than the homogenous mixing models Lockdown has dramatically slowed down the spread of COVID-19 in UK, and even more significantly, has changed the growth in the total number of infected from exponential to quadratic. This significant change is due to a transition from a mobility-driven epidemic spreading to a spatial epidemic which is dominated by slow growth of spatially isolated clusters of infected population. Our results strongly indicate that, to avoid a return to exponential growth of COVID-19 (also known as “second wave”), mobility restrictions should not be prematurely lifted. Instead mobility should be kept restricted while new measures, such as wearing of masks and contact tracing, get implemented in order to prevent health services becoming overwhelmed due to a resurgence of exponential growth.

## 1. Introduction

The UK government announced on 23 March 2020 full lock-down measures aimed at decelerating the spread of COVID-19 in the country. In this report we analyse the officially available data on the spread of COVID-19 in UK, both before and after the lockdown. We show that before lock-down the growth in the total number of infected cases was exponential, with an exponent *α = 0.207*, and argue that this exponential growth is best described with a mobility-driven SIR/SEIR model, e.g. [2], which explicitly incorporates deterministic mobility (e.g. daily commute to work and transport) and physical proximity, rather than the conventional homogeneous mixing models. We show that lockdown has greatly decelerated the growth of the epidemic and, even more significantly, has changed the key spreading mechanism of COVID-19 from a mobility-driven exponential growth to a much slower *quadratic growth*. We argue that quadratic growth is a direct consequence of restriction imposed on “deterministic mobility” of population (i.e. for daily commutes to/from work and transportation) which has broken up infected population into largely isolated spatial clusters. By considering the diffusive growth of such clusters, we arrive at modified SIR/SEIR model, which correctly describes quadratic growth of the number of infected case after the lockdown. Key implications of our findings for design of safe exit strategies are a discussed. We strongly recommend that mobility remains restricted, while other lockdown measures are cautiously lifted. Our findings shed serious doubts on the ability of homogenously mixed SIR/SEIR modelling to correctly guide the design and of such strategies.

The rest of this paper is organised as follows In Section 2 we describe the data sources used in our study and briefly discuss the implications of the limited testing for COVID19 infection on the quality of data. In Section 3 we provide our analysis for the growth of the total number of COVID-19 infected cases, both prior and after the lockdown. We conclude in Section 4 with summary and a discussion of the implications of our finding for UK exit strategies.

## 2. Data sources

We have used data published by the UK Government Public Health England (PHE) and Department of Health and Social Care ^1^as a primarily source for our study and cross-checked this data with a number of publicly available sources. UK Government publishes on a daily basis data on the total number COVID-19 tests carried out, the total and daily number of infected cases as obtained from testing, and the total number of new infected cases.. Due to the stochastic nature of the epidemic, the number of new reported cases show strong statistical fluctuation. On the other hand, the total number of infected cases show a remarkably smooth behaviour. For this reason, in the rest of this paper we will focus our attention primarily on analysing data on the total number of infected cases.

As of 5 May 2020 a total number of 63 667 people were tested in the UK, according to PHE data, which assuming a UK population of 66.5M, corresponds to a sample size of just 1% of population being tested for COVID-19. The testing was initially performed in NHS hospitals and PHE labs on those with clinical needs. It was extended later to health and care workers and their families, and more recently to general population (trough a national surveillance program).^2^

The focus of our study is primarily, on understanding general trends and patterns of COVID-19 spreading growth in UK, rather than absolute growth numbers. Therefore, we expect that our findings are not strongly affected by the current limited availability of data for UK. We will provide a more in-depth analysis of UK’s COVID19 testing strategy and its implications elsewhere.

## 3. Data analysis and models

### a. Analysis and model before lockdown

The standard model of epidemic spreading with immunity or death (or both) from the dieses are the Susceptible-Infected-Removed (SIR) models and its variant, the Susceptible-Exposed-Infected-Removed (SEIR) model [1]. These models assume a homogeneous mixing among population (an infected member of population can infect any other member of population), and in their deterministic form are described by a set of three (four in cases of SEIR) coupled differential equations. These equations describe time evolution of the ratio of the susceptible individuals, S(t), Infected individuals I(t), Exposed individuals E (t), and Removed Individuals, R(t), respectively, in population, as result of epidemic spreading. Removed individuals are those who become immune to the virus after being infected or died due to infection. The equations are usually initialised assuming a very small fraction of infected who start the epidemic, with the rest of population being susceptible to the epidemic (which is the cases for new epidemics such as COVID-19).

Starting from the above infection scenario at time, *t*_0_, the SIR/SEIR model predicts an exponentially fast initial spreading of the disease in the population, with *I(t*) obtained from:

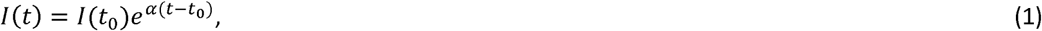

Here, *α* is the growth exponent which depends on the viral infectivity of the epidemics, *λ*, the average number of contacts per unit time, 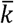, and the rate of recovery from the virus, *δ* and is given by:

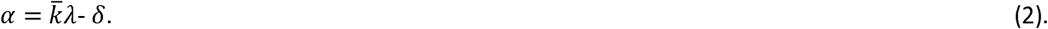

Hence for 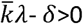 the epidemics spreads exponentially fast starting from few infected, while for 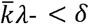 it dies off exponentially fast. We note that 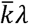 corresponds to the so-called *R*_0_ parameter.

At a later stage, the exponential growth in the number of infected, as obtained from the SIR/SEIR models, is counter-acted by their decay due to the recovery and death mechanisms. Therefore, even in the absence of any prevention measures the number of infected will eventually die out (for a closed population). Consequently, the total number of infected cases, 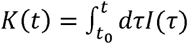 shows an initial exponential growth after which it plateaus, and then stabilises to a final fraction of the population.

Figure 1 (top panel) shows the growth in the total number of known infected cases in the UK starting from 4 March 2020. We founds that an exponential growth of the from:

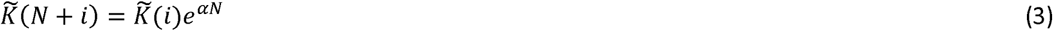

fits the growth data up to 24 March l extremely well. In this figure data points are shown with blue dots while the exponential growth model results are shown with red dots. The growth exponent, was found, from a least-squares fit, to be *α*=0.207.The excellent fit to exponential growth can also be seen from the lower panel in Figure 1, where we have plotted log (*K*) and 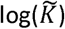 againts time.

**Figure 1.**
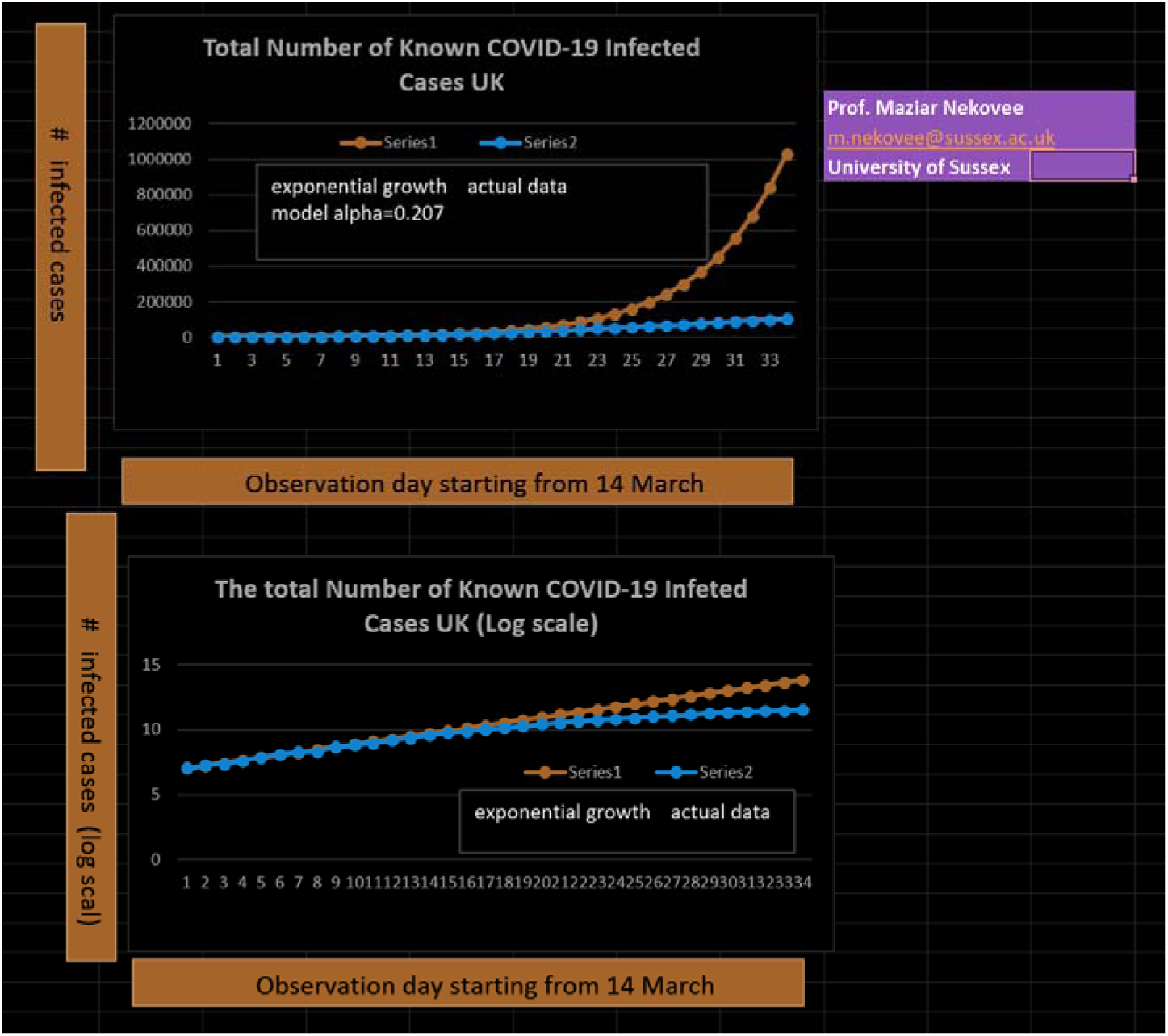
*Growth in the total number of known infected cases in UK, K(t) starting from 14 March 2020 is shown in top panel. Blue dots show actual data and red dots the exponential growth model with exponent α = 0.207. Bottom panel display results shown in log scale*.

COVID-19 is an airborne disease, and its main transmission routes are respiratory droplets (cough, sneeze, heavy breathing) and close contacts [3]. Infected individual “carry” the virus with them and can infect healthy individuals when they are in close contact with them, hence mobility and physical proximity and contact duration are key mechanism for the spread of COVID-19 but these mechanisms are not explicitly captured in the standard SIR/SEIR homogeneous mixing models.

We believe that a model initially proposed by Rhodes and Nekovee [2], in the context of the spreading of Bluetooth viruses among smartphones carried by mobile users, better captures the above key mechanism of COVID-19 spreading In this model individuals move about with deterministic velocities across a metropolitan areas (we call this purposeful mobility, i.e. for work-home commute) as opposed to random mobility. Infected individuals transmit the disses to healthy individuals which are within their infection radius, R, during contacts made in the course of their daily mobility. Consequently, the rate of change in the number of infected individuals as a result of mobility-driven encounters is found to follow the following equation [2]:

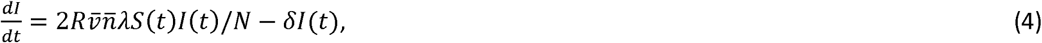

In the above equation 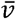 and 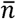 are the average population velocity and density, respectively, and, *R*, is the infection radius. Therefore, unlike the conventional SIR models, the rate of infection is explicitly given in terms of the radius of infection, *R*, population mobility patterns and population density. Furthermore, the model also predicts an initial exponential growth of the epidemic. However, the growth exponent, *α*, depends explicitly on the population mobility, the radius of infection, and population density. And is given by [2]:

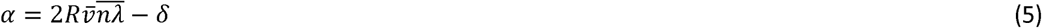

Therefore, the effect of containment mechanisms, such as restricting mobility, enforcing social distancing (to reduce population density) and adjusting infection radius (e.g. by compulsory wearing of masks) can be directly incorporated in the model. We note that, in addition to purposeful mobility, individuals also move randomly and this adds a small component to 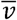 in the above equation, such that in the absence of purposeful mobility (e.. due to lockdown), i.e. with 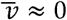, we would expect a drastic slowing down of the epidemic growth, although a slow growth will persist, through a spatial diffusion process, as we discuss next.

### b. Analysis and model after lockdown

UK Government announced its full lock-down rules on 23 March 2020 (A partial lockdown of pubs, restaurant and public gatherings in the UK were announced on 14 March 2020). The lockdown rules stipulate strict constraints on mobility of UK citizens, social mixing, running of business and public gatherings. People are allowed to leave their home only for the below reasons:

- Shopping for basic necessities, which must be as infrequent as possible.
- One form of exercise a day, alone or with members of one’s household.
- any medical need
- Travelling for work purposes, but only where you cannot work from home.

To reduce social contact, the UK Government also ordered many businesses (such as pubs and bars, restaurants), and public venues and gatherings as well as schools to be closed. The lockdown imposed strong constraint of citizen’s regular mobility patterns, to-and-from work, as well as mobility across the country. It also greatly reduces any form of social mixing outside one’s own household (e.g. in parks) during which infection can spread. Furthermore, UK supermarkets and shops have implemented social distancing measures in order to reduce the chance of infective contacts between people during their regular, albeit infrequent (potentially weekly) shopping trips.

In a homogenously mixed SIR/SEIR model, the effect of lockdown measures would translate into a smaller and potentially less infective (due to two meter distancing rule) number of contras per day between infected and susceptible population. The nett effect of lockdown in the SIR/SEIR model is a reduction in that infectivity, *λ*, and the average number of contacts per unit time, k □, and hence the growth exponent *α*, say to 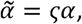 wih 0 < *ҫ* < 1. However, according to the SIR/SEIR model, the epidemic would still grow exponentially after lockdown, albeit with a much smaller exponents 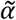.

Figure 2 (top panel, red dots) shows the growth in the total number of infected cases staring from 24 March 2020 up to 24 April 24 2020, i.e. during the first month of lockdown.

Our analysis of the above data shows that an exponential growth model, even with a growth exponent much smaller than the pre-lockdown value, is a poor fit to the data. In contrast, the data for this period fits extremely well a one-parameter quadratic growth (blue dots in Figure 2) of the form:

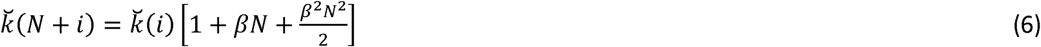

**Figure 2.**
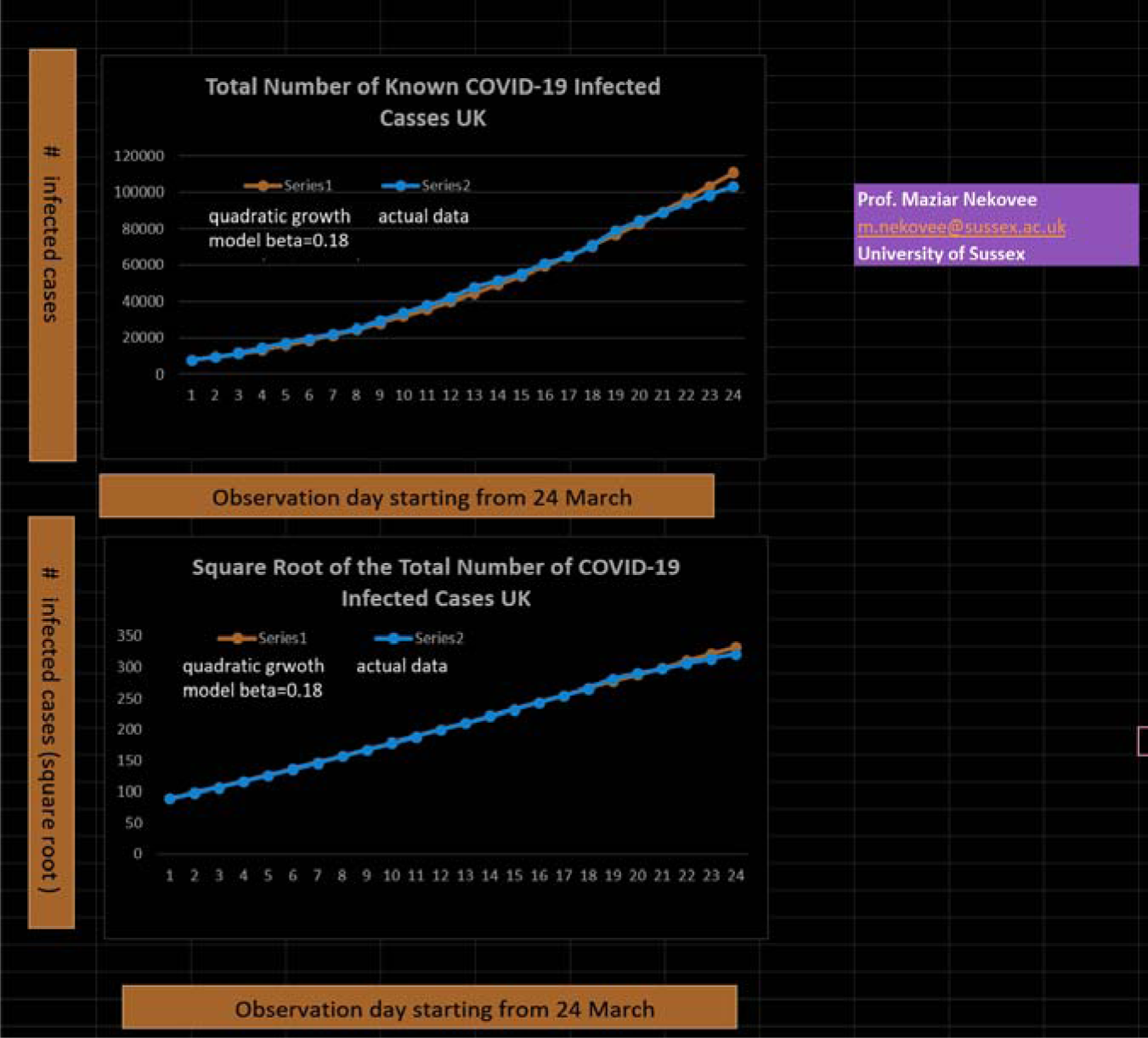
*Growth in the total number of known infected cases in UK, K(t) is shown (top) from 24 March to 24 April. Blue dots are actual data and red dots results of the quadratic growth mode, 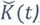, with coefficient β* = 0.18. *Bottom panel shows 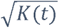 and 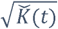*.

With *β* = 0.18. The fitting parameter, *β*, was obatined from a least-squares fit. To further illustrate this finding we display in Figure 2 (bottom) 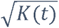 and 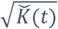 for the above observation window. It can be seen that both these quantise show a clear linear growth with time, further indicating that growth after lockdown has become quadratic, rather than remaining exponential. Quadratic growth has been also observed in analysis of COVID-19 spreading in China [4] but showing up less strongly/clearly than in the UK data.

### c. Understanding the quadratic growth

Figure 3 shows transport use change in UK since the start of lock-down, indicating that daily mobility of UK population is very significantly reduced due to lock-down. Given, also that an exponential model does not fit deceleration of epidemic after lock-down, we conclude that by severely restricting mobility, lock-down has quantitatively changed the spreading mechanism by isolating the infected population into spatial clusters with very limited inter-cluster mobility.

**Figure 3.**
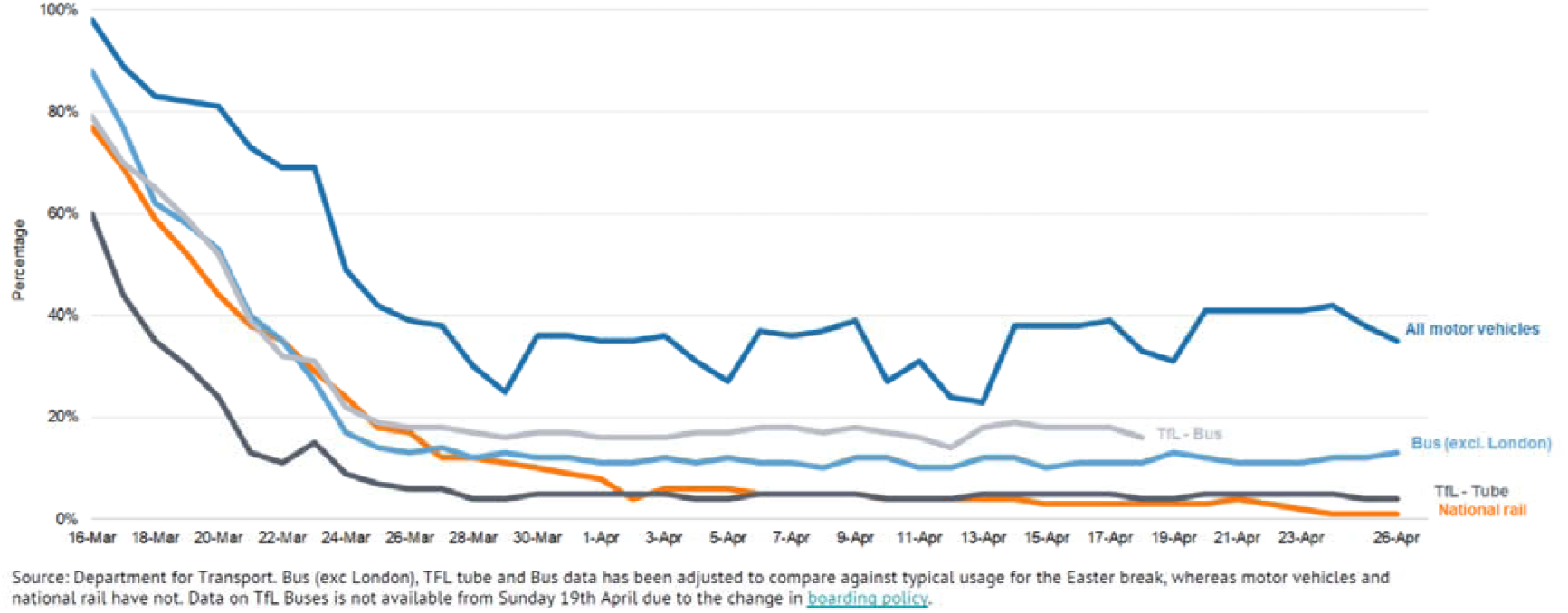
*Transport use change in UK since lock-down. [Source UK Government Daily Briefing on COVID-19, 28/04/2020^3^]*.

We postulate that with limited purposeful mobility across different boroughs of large cities such as London, i.e. limited daily home-office commutes, the epidemic mainly spreads as a result of diffusive growth of these spatial clusters. Similarly lockdown has greatly restrict travel across the UK and hence fragmented the country into spatially isolated clusters of infection.

The rate of increase in the number of infected case resulting from the spatial growth of such a cluster can be described with the following equation:

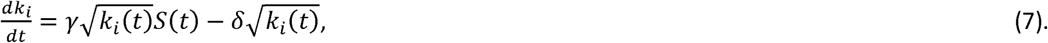

Which, after linearizing has a solution of the quadratic form:

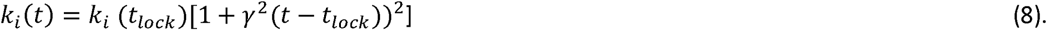

Where *t_lock_* is the onset of lockout and ***γ*** is a parameter which is related to diffusive mobility of population across the cluster.

Alternatively, quadratic growth can be understood from the perspective of graph theory (or network epidemics), where due to lock-down the contact network is changed from a, to good approximation, random graph to a spatial network, where due to high clustering [4], the branching process which that give rise to exponential growth is decelerated to only quadratic growth.

## 4. Summary and implications for exit strategy

Our analysis shows that prior to lockdown the spread of COVID-19 in UK was exponential, with an exponent α=0.207 In case of COVID-19 this spreading patterns is quantitatively better described with mobility-driven SIR-SEIR model [2] rather than the homogenous mixing models Lockdown has dramatically slowed down the spread of COVID-19 in UK, and even more significantly has changed the growth in the total number of infected from exponential to quadratic, up to observation point of 24 April. This significant change is due a transition from a mobility-driven epidemic spreading to a spatial epidemic which is dominated by slow growth of spatially isolated clusters of infected population. Our results strongly indicate that and premature lifting of mobility restrictions, as part of exit from lockdown, will result in the return of COVID-19’s exponential growth, i.e. a second wave of epidemic, and this is illustrated in Figure 4. Instead mobility should be kept restricted while other lockdown measure get eased *in order to prevent health services becoming overwhelmed due to a resurgence of exponential growth*.

**Figure 4.**
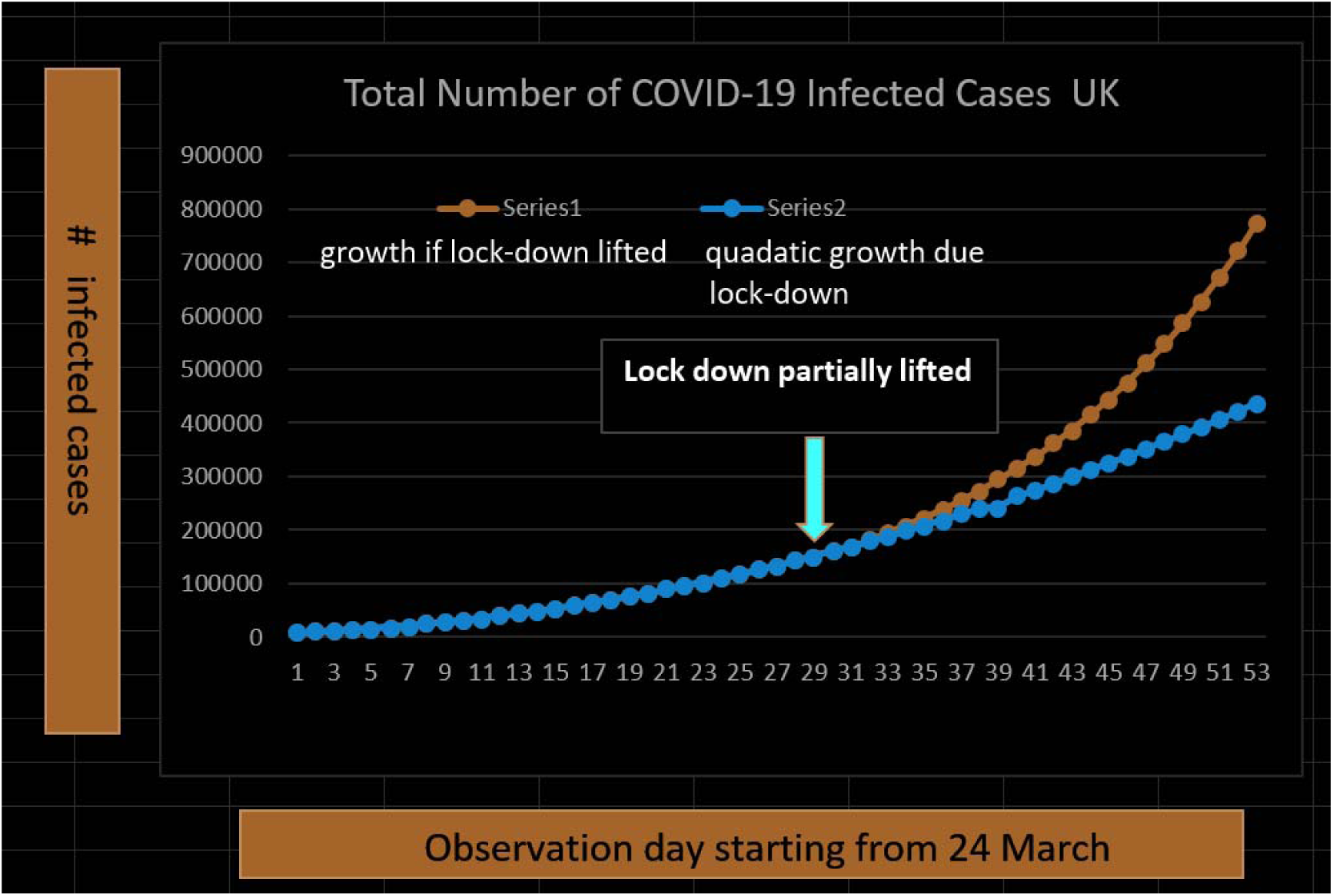
*the figure shows a hypothetical scenario in which mobility restriction measures of lock-down were lifted on 24 April 2020 while other measures such as social distancing are still in place*.

## Data Availability

All data used is publicly available.

https://www.gov.uk/guidance/coronavirus-covid-19-information-for-the-public

## Acknowledgments

The author dedicates this work to the memory of his dear friend Dr Babak Pourbohloul. He thanks Prof. John E. Inglefield and Dr Mona Ghasemian for reading the paper and their helpful comments.

This research was partially funded by the Economic and Social Research Council (ESRC), UK through an Impact Accelerator Account Project “Co-creating Connected and Intelligent Care Homes for People with Dementia”

## Footnotes

1 https://www.gov.uk/guidance/coronavirus-covid-19-information-for-the-public

2 See the UK government’s national testing strategy for more information on COVID-19 testing program, https://www.gov.uk/government/publications/coronavirus-covid-19-scaling-up-testing-programmes

3 https://www.gov.uk/government/collections/slides-and-datasets-to-accompany-coronavirus-pressconferences

